# SARS-CoV-2 seroprevalence in pregnant women during the first three COVID-19 waves in The Gambia

**DOI:** 10.1101/2023.06.09.23291201

**Authors:** Ramatoulie E. Janha, Alasana Bah, Hawanatu Jah, Fatima Touray, Yahaya Idris, Saikou Keita, Yassin Gaye, Samba Jallow, Tisbeh Faye-Joof, Baboucarr Njie, Rachel Craik, Nuredin I. Mohammed, Peter von Dadelszen, Umberto D’Alessandro, Anna Roca, the PRECISE Network

## Abstract

**Objectives:** SARS-CoV-2 transmission in Sub-Saharan Africa has probably been underestimated. Population-based seroprevalence studies are needed to determine the extent of transmission in the continent.

**Methods:** Blood samples from a cohort of Gambian pregnant women were tested for SARS-CoV-2 total IgM/IgG before (*Pre-pandemic1:* October-December 2019 and *Pre-pandemic2:* February-June 2020) and during the pandemic (*Post-wave1*: October-December 2020, *Post-wave2:* May-June 2021; and *Post-wave3*: October-December 2021). Samples positive for total SARS-CoV-2 IgM/IgG were tested for protein-specific antibodies.

**Results:** SARS-CoV-2 total IgM/IgG seroprevalence was 0.9% 95%CI (0.2, 4.9) in *Pre-pandemic1*; 4.1% (1.4, 11.4) in *Pre-pandemic2*; 31.1% (25.2, 37.7) in *Post-wave1*; 62.5% (55.8, 68.8) in *Post-wave2* and 90.0% (85.1, 93.5) in *Post-wave3.* S-protein IgG and NCP-protein IgG seroprevalence also increased at each *Post-wave* period. Although S-protein IgG and NCP-protein IgG seroprevalence was similar at *Post-wave1*, S-protein IgG seroprevalence was higher at *Post-wave2* and *Post-wave3*, [prevalence difference (PD) 13.5 (0.1, 26.8) and prevalence ratio (PR) 1.5 (1.0, 2.3) in *Post-wave2*; and 22.9 (9.2, 36.6) and 1.4 (1.1, 1.8) in *Post-wave3* respectively, p<0.001].

**Conclusion:** SARS-CoV-2 transmission in The Gambia during the first three COVID-19 waves was high, differing significantly from official numbers of COVID-19 cases reported. Our findings are important for policy makers in managing the near-endemic COVID-19.

**Highlights:**

- High specificity of the IgM/IgG SARS-CoV-2 test using samples collected prepandemic
- Very high (>90%) SARS-CoV-2 seroprevalence after third COVID-19 wave in The Gambia
- High SARS-CoV-2 transmission contrasts with low number of COVID-19 reported cases

## INTRODUCTION

SARS-CoV-2, the causative agent of the coronavirus disease 2019 (COVID-19), was first identified in December 2019 in Wuhan, China [1]. SARS-CoV-2 was rapidly transmitted globally, disrupting structures and systems; and causing millions of deaths. Globally, there were 278.7M COVID-19 cases and 5.39M attributable deaths by December 2021 [2], 21 months after the World Health Organization (WHO) declared COVID-19 a pandemic [3]. Nigeria reported the first case in Sub-Saharan Africa (SSA) on 28 January 2020 [4]. The SSA continent had the worst epidemiological modelling predictions of cases, hospitalisations and deaths [5]. SARS-CoV-2 infection during pregnancy is of particular concern, as it increases the rates of adverse maternal and neonatal outcomes [6]. Nevertheless, the impact of COVID-19 in SSA was lower than expected, with 7.1 million cases and 155,300 deaths by December 2021, i.e., 2.5% of all cases and 2.9% of all deaths worlwide, despite Africa representing 17.4% of the global population [2].

Some underpinning factors in Africa, specifically in SSA, may explain the discrepancy between predictions and actual burden. One of these is the limited capacity for diagnosis, surveillance and testing for SARS-CoV-2, exacerbated by lack of systematic death registrations in most SSA countries. Moreover, older age is a major risk factor for severe COVID-19 and associated deaths. As the SSA population is young, i.e., in 2022, the median age was 18.8 years, with 40% aged 14 years and younger, this may partially explain the low number of reported deaths [7]. The communities’ perceptions of COVID-19 and avoidance of healthcare facilities [8] during the pandemic may have added to underdiagnosis of cases and non-certification of COVID-19 as the cause of death. However, there is emerging evidence that transmission in SSA has been at least as high or even higher than in other continents [9], although these data should be interpreted with caution as some studies have important limitations. Most of them do not represent the entire population, rather specific groups, generally high risk groups [10]. In addition, SARS-CoV-2 prevalence may be overestimated as SARS-CoV-2 antibodies may cross-react with other viral or parasitic antibodies in the region, such as IgG antibodies against the spike and nucleocapsid proteins of different human coronaviruses: HCoV-OC43, HKU-1, NL63 and 229E, as well as SARS and MERS [11], and IgG directed against *Plasmodium falciparum* antigens where malaria is endemic [12]. To overcome such challenges, the WHO recommends: (i) the use of multiple assays to confirm seropositivity for SARS-CoV-2, including testing positive or equivocal samples with neutralizing assays; (ii) repeated cross-sectional surveys to monitor the population serological profile [13]; and (iii) testing assay specificity [14], as the multiple available assays consist of a combination of serological tests targeting separate groups or monomers of specific antigens.

The Gambia is a small country situated in West Africa with a population of 2.57 million [15]. The objective of this study was to describe the seroprevalence of SARS-CoV-2 before (as a measure of specificity) and following each of the first three waves among pregnant women living in peri-urban and rural Gambia who participated in the PRECISE study [16]. In general, pregnant women represent a healthy young adult population and seroprevalence in this group should represent seroprevalence in adult healthy population.

## METHODS

### COVID-19 and the National response in The Gambia

The Gambia’s first official COVID-19 case was reported on 16 March 2020 [17]. By December 2021, The Gambia had registered three SARS-CoV-2 transmission waves dated: first, July-September 2020; second, January 2021; and third, July 2021 (Figure 1). The first and third waves were more intense, with higher peaks of official daily cases [18] with the delta variant being the most frequent during the third wave [19]. The second wave concurred with the global transmission of the delta/B.1.617.2 subtype variant, described in The Gambia at the time [19].

**Figure 1:**
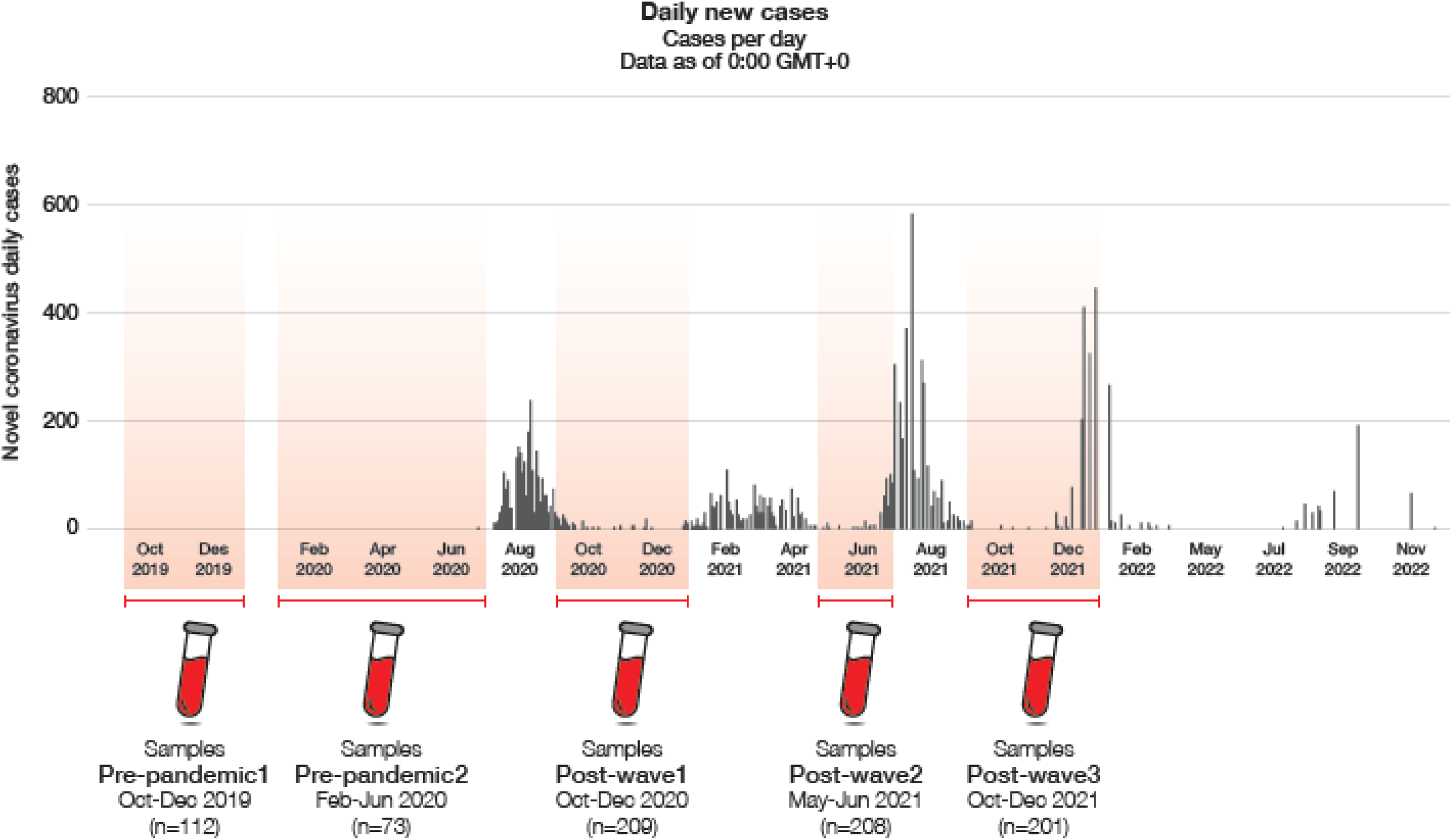
SARS-CoV-2 transmission waves embedding study design. A schematic design showing sampling time periods and sample-sizes per time period. The shadowed areas are the period of sample collection. Samples were grouped into two *Pre-pandemic* and three pandemic time periods linked to the first three COVID-19 waves as *Post-wave* time periods.

Following the diagnosis of the first few official cases, a state of emergency was declared [20]. Briefly, authorities prompted stringent response measures, including border closures and a national lockdown (schools, worshipping grounds, and non-essential business activities), initially for three months. Mandatory wearing of face masks and observing physical distancing in public places were enforced [20]. Epidemiological surveillance consisted of isolation, quarantine and Rt-PCR testing of suspected cases and travellers, and contacts-tracing of confirmed cases. Surveillance, though, happened mostly carried out in the urban Western Region. Vaccination coverage was very low; by December 2021, only 10.2% of the total population (and <1% in our study area population) was fully vaccinated with 2 doses of Astrazeneca/Sinopharm or 1 dose of Johnson and Johnson [21].

### Study design – The PRECISE study

This is an ancillary study of the PRECISE study (Pregnancy Care Integrating Translational Science, Everywhere) [16], in which a pregnancy cohort was recruited in The Gambia, Kenya and Mozambique, with the collection of extensive sociological and clinical data and biological samples throughout woman’s pregnancy, until 6 weeks post delivery [16].

### PRECISE The Gambia

In the Gambia, the study was implemented in Farafenni, North Bank Region, 116 km from the capital city Banjul. Farafenni is divided into rural and urban areas and forms an important commercial border with Senegal. Study participants were pregnant women attending antenatal care at three health facilities: Maternal Newborn Child and Adolescent Health Clinic in Farafenni (urban), Illiasa and Ngayen Sanjal (rural) health centres. Farafenni General Hospital is the main referral hospital for the North Bank Region, population 225,516 [22], and Illiasa and Ngayen Sanjal are 17km west and 20km east of Farafenni, respectively.

The Gambian PRECISE cohort includes 1,253 pregnant women aged 16-49 years. Participants were recruited at their first antenatal visit which could be at any time during their pregnancy (mainly in 2^nd^ trimester). A second antenatal visit was scheduled where possible between 28 weeks’ gestation and the onset of labour (in 3^rd^ trimester), and at least 4 weeks after their booking visit. Women were then seen during their admission for giving birth if that occurred in a participating healthcare centre. The last study visits occurred 6-weeks to 6-months after delivery. At each study visit, blood samples were collected (between October 2019 and March 2022). As stated, demographic, epidemiological and clinical information was collected during study visits. In our study population, vaccination status was not collected.

### SARS-CoV-2 sero-prevalence study

For the SARS-CoV-2 seroprevalence study presented here, we considered five sera collection time periods that corresponded to the SARS-CoV-2 waves observed in The Gambia [19]: (i) *Pre-pandemic1* (October - December 2019) before any case was detected in The Gambia; (ii) *Pre-pandemic2* (February – June 2020) during which few cases were diagnosed but no wave had yet started and the stringent 3-months national lockdown implemented; (iii) *Post-wave1* (October – December 2020); *Post-wave2* (May – June 2021); and (v) *Post-wave3* (October – December 2021) (Figure 1). The *Pre-pandemic1* period was included as an additional step to determine specificity of the test used to measure the study primary endpoint in our population as no SARS-CoV-2 transmission in The Gambia during these months is expected.

For each time period, only one sample per woman was included although samples from the same woman could have been selected for other periods.

### Study endpoints

The primary endpoint was the SARS-CoV-2 seroprevalence (IgM and IgG against all SARS-CoV-2 proteins) at each time-period as determined by the WANTAI. Secondary endpoints include IgG seroprevalence to specific SARS-CoV-2 proteins (spike S1-subunit IgG and nucleocapsid protein, NCP-IgG) (EUROIMMUN test).

### Serum collection and preparation

Venous blood (6ml) was collected into serum separation tubes (BD Vacutainer SST^TM^ tubes, Fisher Scientific), allowed to clot, and centrifuged at 2000g for 15 minutes. Separated sera were kept at -80°C until serological analyses.

### Lab analysis and interpretation

a. **WANTAI immunoassay.** Sera were tested using the WANTAI SARS-CoV-2 Ab ELISA Rapid Diagnostic Test kit (Wantai, Beijing, China) for qualitative detection of total IgM and IgG against SARS-CoV-2 [23]. The WANTAI immunoassay microwell strips are pre-coated with recombinant receptor-binding domain of the spike protein antigen incorporated into a lateral flow ELISA design. The assay has a manufacturer-validated sensitivity of 98.7% and specificity of 98.6% in participants from China [23]. An independent clinical agreement validation of the WANTAI assay was conducted at the Frederick National Laboratory for Cancer Research in the United States in July 2020 [23]. Each microwell plate had a blank calibrator, 50µl control, l00µl of serum. Following two-steps of incubation at 37°C, washing, horse radish peroxidase-conjugation and colourisation, absorbance was read at 450nm. After blanking and cut-off determination, samples with absorbance/cut-off ratio ≥1 were positive for WANTAI whole IgM/IgG for SARS-CoV-2.
b. **EUROIMMUN immunoassay.** Two EUROIMMUN anti-SARS-CoV-2 ELISA test kits (EUROIMMUN, US, Germany) [24, 25] were further performed on all WANTAI tested-positive samples (Supplementary Figure 1). The EUROIMMUN microplates are pre-coated separately with recombinant S1 domain of spike protein, for detecting IgG antibodies against the S1-subunit; and modified NCP for detecting IgG against NCP. The S1-IgG test has a sensitivity of 80% and a specificity of 98.5% [24]; and the NCP-IgG test has a sensitivity of 94.6% and a specificity of 99.8% on European patients’ sera [25]. Samples were diluted in sample buffer. Each well had 100µl of calibrators. Plates were incubated, washed and enzyme-conjugated, followed by absorbance measurement at 450nm. Results for controls and samples were calculated considering the extinction of the calibrator. The results were interpreted as: ratio<0.8: negative; ratio ≥0.8 to <1.1: borderline/indeterminate; and ratio ≥1.1: positive.

### Statistical Analysis

Categorical variables, including SARS-CoV-2 seroprevalence, were summarized using proportions. Chi-square or Fisher’s exact test were used where appropriate to test for differences in proportions between time periods. Continuous variables were summarized using median (interquartile range).

SARS-CoV-2 seroprevalence by WANTAI was expressed as the proportion [95%CI] of WANTAI-positives to the total number of tested samples per time period. SARS-CoV-2 seroprevalence by EUROIMMUN was expressed as the proportion [95%CI] of EUROIMMUN-positives to the total number of samples tested per time period. EUROIMMUN borderline results were considered as negatives.

Prevalence ratios (PR) and prevalence differences (PD) between EUROIMMUN S-IgG and NCP-IgG antibodies at each wave were calculated using generalized linear models (glm) with robust standard errors. We used Bonferroni method throughout to account for multiple comparisons (due to multiple time periods).

To determine the proportions of new infections at each post-wave period (considering any seroprevalence by WANTAI), our assumptions were that after one infection women remained seropositive throughout the study and women tested at each time period represented a random sample from the PRECISE cohort. Therefore, we calculated the percentage of new infections as the increased prevalence in the population at risk (percentage of population uninfected in the preceding wave). All data management and analyses for this study were performed using Stata version 17.0 [26].

## RESULTS

### Participants’ baseline characteristics

803 women were included in the five sera collection time periods, with 112 in *Pre-pandemic1*, 73 in *Pre-pandemic2*, 209 in *Post-wave1*, 208 in *Post-wave2* and 201 in *Post-wave3*. Baseline characteristics of women were similar at the different time points, with a median age of 26 years, both Mandinkas and Wolofs being the most common ethnic groups, median household size of 9 individuals per household and median parity 2-4 children per woman (Supplementary Table 1).

### SARS-CoV-2 sero-prevalence (WANTAI)

Overall, SARS-CoV-2 seroprevalence was low in the *Pre-pandemic1 period* at 0.9% 95%CI (0.2, 4.9) and the *Pre-pandemic2 period* at 4.1% (1.4, 11.4) (Table 1a, Figure 2a). SARS-CoV-2 seroprevalence significantly increased over time, from 31.1% (25.2, 37.7) in the *Post-wave1 period* to 90.0% (85.1, 93.5) in the *Post-wave3 period* (p<0.001, Figure 2a, Table 1a and Table 1b). The stratified analysis of seroprevalence shows similar results for urban and rural Farafenni (Table 1a, Figure 2b).

**Table 1a.** Overall SARS-CoV-2 seroprevalence (IgM and IgG combined) at each study period.

| Area | Period | Positive (Total)<br>n (N) | Prevalence (95% CI) |
| --- | --- | --- | --- |
| **Overall** | Pre-Pandemic1 | 1 (112) | 0.9 (0.2, 4.9) |
|  | Pre-Pandemic2 | 3 (73) | 4.1 (1.4, 11.4) |
|  | Post-Wave1 | 65 (209) | 31.1 (25.2, 37.7) |
|  | Post-Wave2 | 130 (208) | 62.5 (55.8, 68.8) |
|  | Post-Wave3 | 181 (201) | 90 (85.1, 93.5) |
| **Urban** | Pre-Pandemic1 | 1 (87) | 1.1 (0.2, 6.2) |
|  | Pre-Pandemic2 | 3 (68) | 4.4 (1.5, 12.2) |
|  | Post-Wave1 | 23 (77) | 29.9 (20.8, 40.8) |
|  | Post-Wave2 | 64 (97) | 66 (56.1, 74.6) |
|  | Post-Wave3 | 68 (75) | 90.7 (82, 95.4) |
| **Rural** | Pre-Pandemic1 | 0 (25) | 0 |
|  | Pre-Pandemic2 | 0 (5) | 0 |
|  | Post-Wave1 | 42 (132) | 31.8 (24.5, 40.2) |
|  | Post-Wave2 | 66 (111) | 59.5 (50.2, 68.1) |
|  | Post-Wave3 | 113 (126) | 89.7 (83.1, 93.9) |

**Table 1b.** Comparison of SARS-CoV-2 seroprevalence (IgM and IgG combined) between post-wave time-periods.

| Name | Period | Estimate (95% CI) | P-value |
| --- | --- | --- | --- |
| **Prevalence Ratio** |  |  |  |
| WANTAI<br>SARS-CoV-2 | PostWave2 vs PostWave1 | 2 (1.5, 2.7) | <0.001 |
|  | PostWave3 vs PostWave1 | 2.9 (2.2, 3.7) | <0.001 |
|  | PostWave3 vs PostWave2 | 1.4 (1.3, 1.7) | <0.001 |
| **Prevalence Difference** |  |  |  |
| WANTAI<br>SARS-CoV-2 | PostWave2 vs PostWave1 | 31.4 (20.3, 42.5) | <0.001 |
|  | PostWave3 vs PostWave1 | 58.9 (49.8, 68.1) | <0.001 |
|  | PostWave3 vs PostWave2 | 27.5 (18, 37.1) | <0.001 |

**Figure 2:**
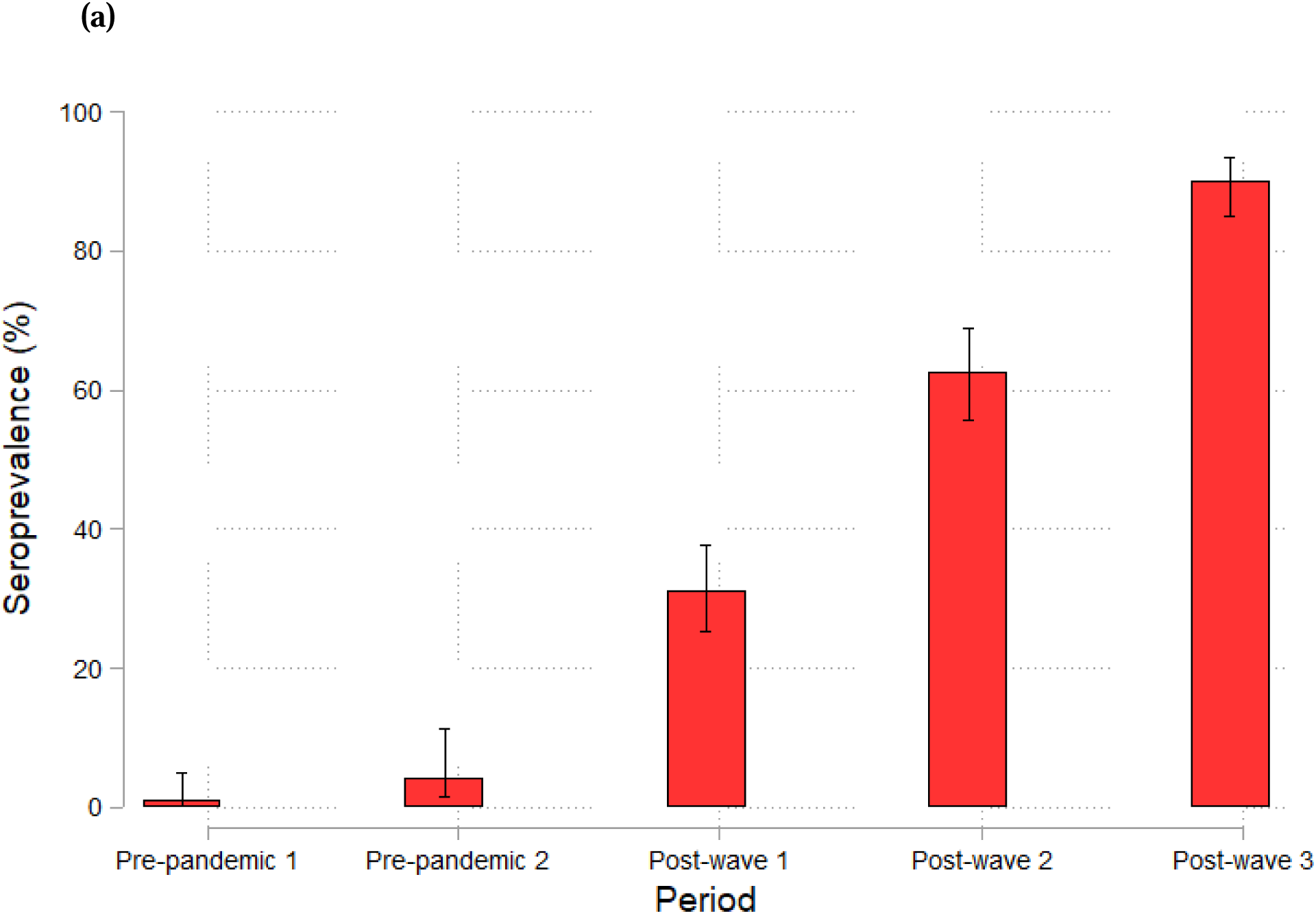
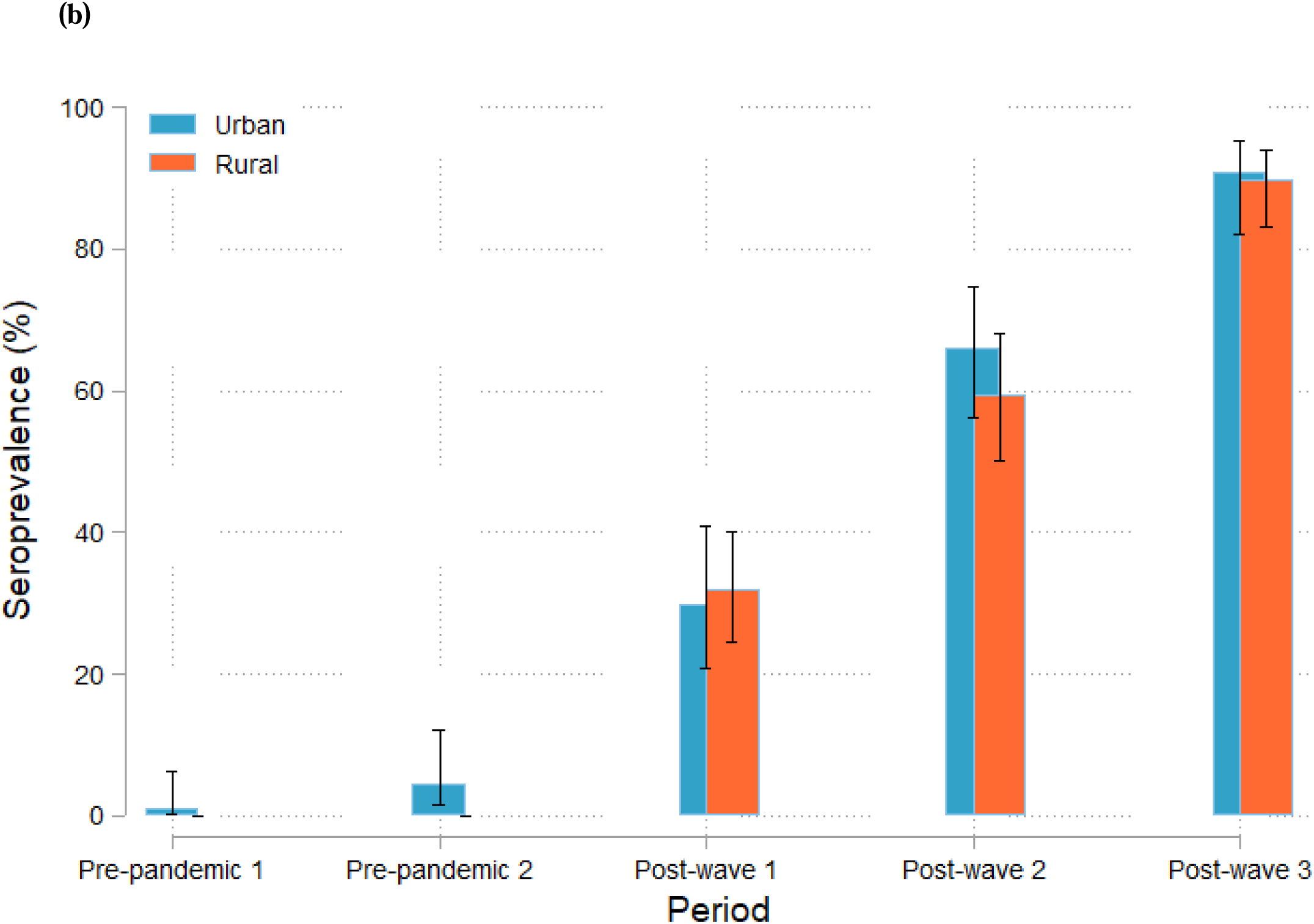
SARS-CoV-2 seroprevalence at each study period: (a) Overall, (b) Stratified by rural and urban health facilities (WANTAI Test). Chart showing SARS-CoV-2 total IgG/IgM antibodies’ seroprevalence with 95% CI (a) per time period and (b) for rural and urban Farafenni in the *Post-wave* time periods

### SARS-CoV-2 protein specific sero-prevalence: Spike (S1-subunit protein) specific IgG and nucleocapsid -NCP protein specific IgG

S-protein specific SARS-CoV-2 sero-prevalence increased significantly in the post-wave periods, from 16.3% (11.9, 21.9) in the *Post-wave1 period* to 74.6% (68.2, 80.1) in *the Post-wave3 period*. Increase of sero-prevalence between *Post-wave1*, *-2* and *-3* was significant for all comparisons, in terms of PR and PD (Supplementary Table 2, Figure 3).

**Figure 3:**
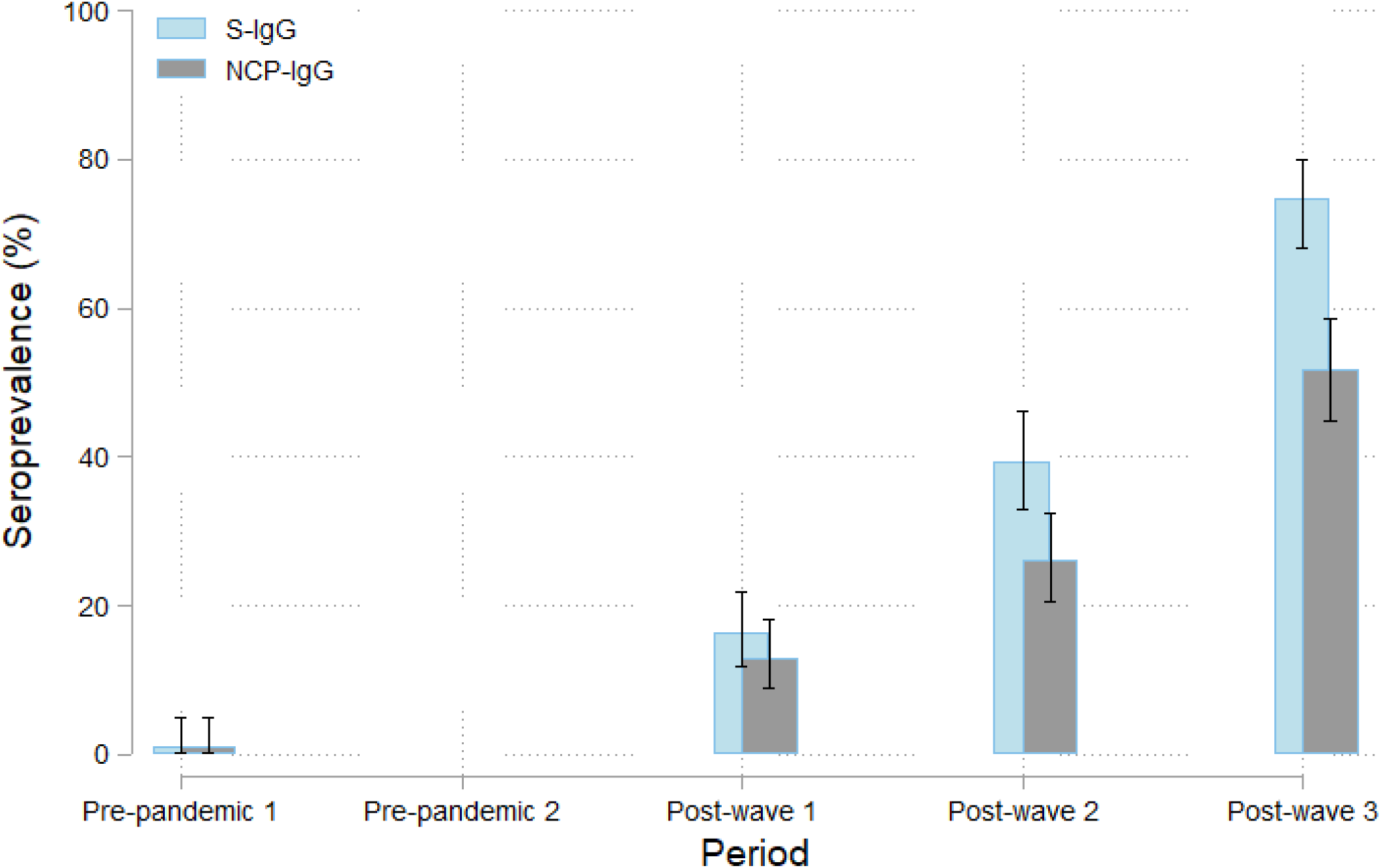
Prevalence of SARS-CoV-2 IgG sero-positivity against S-specific protein IgG and NCP-specific protein IgG. Chart showing SARS-CoV-2 spike S1-subunit protein specific [S-IgG (blue)] and nucleocapsid protein specific [NCP-IgG (grey)] antibodies’ prevalence and 95% CI per time period.

Similarly, NCP-protein specific SARS-CoV-2 sero-prevalence increased significantly in the *post-wave* periods, from 12.9% (9.0, 18.1) in the *Post-wave1 period* to 51.7% (44.9, 58.6) in the *Post-wave3 period*, with significant increase for each comparisson, in terms of PR and PD (Supplementary Table 2, Figure 3).

SARS-CoV-2 protein specific sero-prevalence was similar for S-protein IgG and NCP-protein IgG in the *Post-wave1 period* (p=1.000 for PR and PD). However, SARS-CoV-2 protein specific seroprevalence was higher for S-protein IgG than NCP-protein IgG in the *Post-wave2 period* PR 1.5 95%CI (1, 2.3), p=0.061, PD 13.5 95%CI (0.1, 26.8), p=0.047 and the *Post-wave3 period* PR 1.4 95%CI (1.1, 1.8), p<0.001, PD 22.9 95%CI (9.2, 36.6), p<0.001, showing that the difference between S-protein IgG and N-protein IgG increases after the *Post-wave1 period* (Supplementary Table 2).

### New infections by post-wave period

During *Post-wave2*, 45.6% of the women at risk were newly infected. This percentage was up to 73.3% in the *Post-wave3* period (Figure 4).

**Figure 4:**
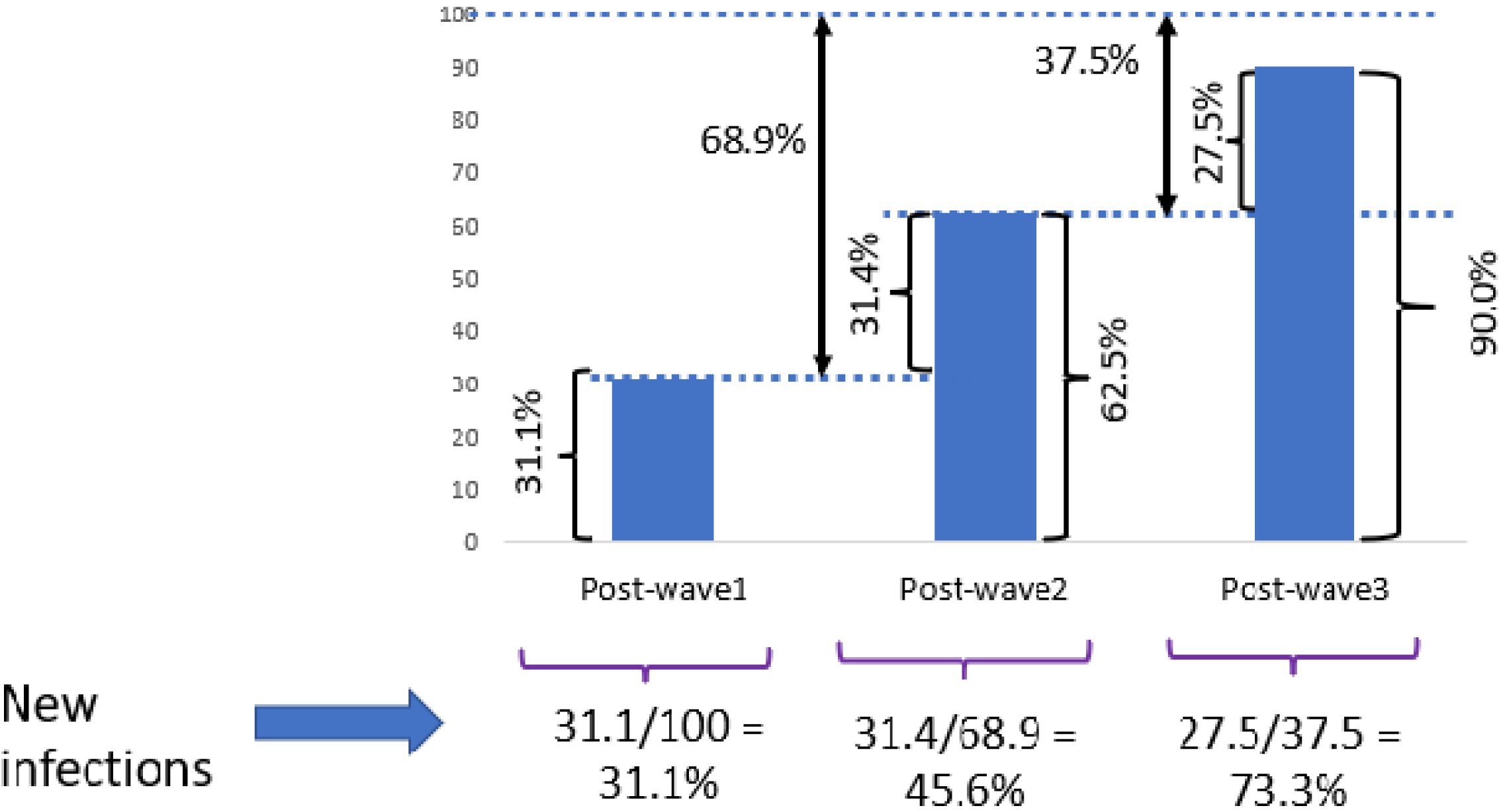
New infections in percentages at each of the *Post-wave* periods. Chart showing percentage of new infections at each *Post-wave* time period.

## DISCUSSION

SARS-CoV-2 sero-prevalence increased significantly among pregnant women living in Farafenni; between October 2019, before the first case of COVID-19 was diagnosed in The Gambia, and December 2021, after 3 epidemic waves. Most women acquired antibodies against SARS-CoV-2, indicating very high transmission in this area, and probably across The Gambia. The discrepancy between such high seroprevalence and the relatively modest number of reported COVID-19 cases is remarkable.

The delayed start of the epidemic in The Gambia was probably due to the measures taken by the Ministry of Health by declaring the state of emergency in March 2020, soon after the first SARS-CoV-2 case was diagnosed in The Gambia [20]. However, the stringent non-pharmaceutical interventions lasted only until June 2020, just before the first wave. Therefore, it is not surprising that in October-December 2020, after the first wave, about a third of pregnant women had SARS-CoV-2 antibodies as adherence to any non-pharmaceutical interventions (e.g., physical distancing, wearing of facemasks, decreasing mass gatherings at public places) from the moment the state of emergency was relaxed and the borders opened was low [17]. These findings confirm that SARS-CoV-2 transmission in The Gambia was much higher than those reported officially about COVID-19 cases and deaths. Indeed, a study analysing the incidence of SARS-CoV-2 after the first wave among staff with standard risk of infection (excluding those working with infected patients) working at the MRC Unit The Gambia indicated that transmission was 80-fold higher than officially reported [20]. Interestingly, the high transmission of SARS-CoV-2 during the first wave did not translate in overall excess mortality, confirming that the epidemic has been milder than in other regions. Nevertheless, in Farafenni, it was detected an excess mortality among the vulnerable 65+ years age-group [27]. Transmission became even easier for the virus in the second and third waves when nationwide restrictions were eased further [17]. The third wave in The Gambia was dominated by the highly transmissible delta variant [19]. This is shown by the estimated 73.3% of the population at risk infected during this wave, increasing the overall SARS-CoV-2 seroprevalence to 90%.

The high SARS-CoV-2 total antibodies’ prevalence described in our study is comparable to that reported from other West African countries at similar times. In Senegal, national SARS-CoV-2 seroprevalence was 28.1% in October-November 2020, at the end of their first transmission wave [28], similar to the prevalence in Farafenni at the same time. Furthermore, in Togo, nationwide seroprevalence in May-June 2021 was 64.3%, which is similar to the prevalence in Farafenni after the second wave [29].

The WHO-procured WANTAI test for standardisation is widely used in Africa [9]. We measured the specificity of the test in our study population by analysing samples collected before the detection of any COVID-19 case outside Wuhan (our *Pre-pandemic1* period covers the period October to December 2019), and were able to confirm the high specificity of this test in our population as only 1 out of 112 samples were positive (likely, a false positive) in the *Pre-pandemic1 period*. A major strength of this study is that pre- and post-epidemic samples were collected from the same population and, therefore, were comparable, and provides confidence that seroprevalence was estimated correctly. This is important as other studies carried out in SSA suggested that the high prevalence of SARS-CoV-2 antibodies in some countries were due to the high cross-reactivity of samples and thus the low specificity of the tests used [11]. Four ELISA tests had lower specificity when used on African samples than European or American samples [30]. These tests included the two EUROIMMUN tests used in this study but not the WANTAI test. We did not determine the specificity of the EUROIMMUN assay as only samples positive by WANTAI were subsequently tested using the SARS-CoV-2 protein specific IgG tests (S-protein and NCP-protein). We also included in our analysis a time period after the first cases had been detected in The Gambia, but before the first wave of the pandemic was officially declared (*Pre-pandemic2*). The seroprevalence in this period did not differ significantly from the *Pre-pandemic1 period*, indicating that transmission was probably low before the first wave was detected even in Farafenni, where the first wave may have started due to the porous border with Senegal [19].

The lower prevalence observed against S-protein IgGs and NCP-protein IgGs compared with whole SARS-CoV-2 IgM/IgG is expected as the latter includes more epitopes and antibody types. Notwithstanding, the IgG prevalence against the S-protein and NCP-protein was high and increasing over time with the occurrence of the COVID-19 waves. Interestingly, the prevalence of NCP-protein IgG is significantly lower than that of the S-protein IgG in the *Post-wave2* period and *Post-wave3 period*, but not in the *Post-wave1 period*. These results suggest antibodies against S-protein are lasting longer than those against the NCP-protein [31], as the latter wane sooner while the former accumulates. Indeed, the first exposure to SARS-CoV-2 of women whose samples were collected in October - December 2020 was during the first wave and thus they had similar levels of S- and NCP-protein specific IgGs. However, during subsequent waves, some women were probably re-infected while others were infected for the first time and thus S-protein specific IgG had a higher prevalence than NCP-protein specific IgG after the second and third waves. An analysis of samples of approximately 39,000 individuals in a clinical laboratory in the United States to monitor SARS-CoV-2 S-protein and NCP-protein durability reveals that both protein-specific IgGs reached a peak of 90% positivity at 21 days post-index [32]. However, by 10 months the NCP-protein IgG seropositivity declined to 68.2%, whilst the S-protein IgG remained at a rate of 87.8%. The data describing longitudinal monitoring of SARS-CoV-2 humoral antibody responses mostly come from high-income countries. Our observation of a shorter durability of NCP-protein IgG than S-protein IgG is in line with those observations.

We need to acknowledge some limitations of our study. The surveys were conducted in a homogeneous population, in a specific region and a relatively narrow age group. Therefore, it is not possible to extrapolate these results to other age groups or regions. However, pregnant women are generally a healthy population who routinely visit hospitals for antenatal care and institutional delivery [33] and, thus, are often used as a proxy group for the whole population. In general, they continue doing similar activities than before pregnancy. Prevalence in rural and urban Farafenni was very similar, possibly suggesting such prevalence may be consistent across the country. In addition, the differences between S-protein specific IgG and NCP-protein specific IgG sero-prevalence in *Post-waves -2* and *-3* may be confounded by vaccination as vaccination only generates antibody responses towards the S-protein. Although vaccination status was not available for study women, it is worth noting that vaccine coverage in the study area was less than 1% by December 2021[21].

In conclusion, SARS-CoV-2 transmission in The Gambia was high during the first three COVID-19 waves. The high seroprevalence observed in our study combined with the former data of no excess mortality in The Gambia in 2020 [27] confirms that the clinical severity of infection during the COVID-19 epidemic has been milder in The Gambia than in other countries. Our results are important for policy makers to take decisions on how to manage the epidemic virus that is progressing into an endemic virus. Our study confirms that in settings with limited laboratory capacity, seroprevalence studies are a powerful tool to understand the real burden of SARS-CoV-2 transmission.

## Data Availability

All data produced in the present study are available upon reasonable request to the authors.

## Authors’ contributions and other declarations

AR conceptualized and designed the study. REJ, UDA and AR interpreted the data and wrote the manuscript. AB and TFJ coordinated the laboratory analysis and edited the manuscript. HJ contributed to conception of the study, acquisition of data, and editing of the manuscript. NIM conducted the data analysis, and contributed to the interpretation of results and editing the manuscript. FT and YI contributed to data acquisition, curation, and editing the manuscript. SK, YG and SJ contributed to data acquisition, analysis and interpretation, and editing of the manuscript. BN contributed to data curation and management, and editing the manuscript. RC and PvD contributed to the study conception, interpretation of data, and editing of the manuscript. The final draft was reviewed and approved by all authors.

## Acknowledgements

We would like to express sincere thanks to the study participants residing in Farafenni. We thank Thomas Mendy for data management oversight and Fatoumata Kongira heading the field and clinical PRECISE team in sensitization and recruitment. We thank all Gambian PRECISE staff for data and sample collection, sample processing and storage. We thank the MRCG at LSHTM platform departments for research support. We are grateful to WHO for providing the WANTAI test kits.

## *The PRECISE Network authorship

King’s College London (Laura A. Magee, Hiten Mistry, Marie-Laure Volvert, Thomas Mendy, Lucilla Poston, Jane Sandall, Rachel Tribe, Sophie Moore, Tatiana Salisbury); Aga Khan University, Nairobi (Marleen Temmerman, Angela Koech, Patricia Okiro, Geoffrey Omuse, Grace Mwashigadi, Consolata Juma, Isaac Mwaniki, Alex Mugo Joseph Mutunga, Moses Mukhanya, Onesmus Wanje, Marvin Ochieng, Emily Mwadime); Centro de Investigação de Saúde de Manhiça (Esperança Sevene, Corssino Tchavana, Salesio Macuacua, Anifa Vala, Helena Boene, Lazaro Quimice, Sonia Maculuve, Eusebio Macete, Inacio Mandomando, Carla Carillho); Donna Russell Consulting (Donna Russell); London School of Hygiene and Tropical Medicine (Hannah Blencowe, Veronique Filippi, Joy Lawn, Matt Silver, Joseph Waiswa, Ursula Gazeley); Midlands State University (Prestige Tatenda Makanga, Liberty Makacha, Reason Mlambo), MRC Unit The Gambia at LSHTM (Andrew M. Prentice, Melisa Martinez-Alvarez, Brahima Diallo, Abdul Sesay, Sambou Suso, Yahaya Idris, Fatoumata Kongira, Modou F.S. Ndure, Lawrence Gibba, Abdoulie Bah, Yorro Bah.); University of Oxford (Alison Noble, Aris Papageorghiou); St George’s, University of London (Judith Cartwright; Guy Whitley, Sanjeev Krishna); University of British Colombia (Marianne Vidler, Jing (Larry) Li, Jeff Bone, Mai-Lei (Maggie) Woo Kinshella, Domena Tu, Ash Sandhu, Kelly Pickerill); Imperial College London (Ben Barratt)

## Ethical Approval

This study was reviewed by the institutional review board, Scientific Coordinating Committee, of the MRCG at LSHTM and ethical approval granted by The Gambia Government/MRCG Joint Ethics Committee and the LSHTM Observational Ethics Committee, ethics ref 22628. All participants provided broad written-informed consent for participation in the PRECISE project and use of their biological samples in future health research. The work described was carried out in accordance with the Declaration of Helsinki for research involving human participants.

---

**Supplementary Figure 1:** Testing algorithm for SARS-CoV-2 seroprevalence by WANTAI and EUROIMMUN Commercial Kits. Chart showing assay work flow. All samples positive for WANTAI test were tested using both EUROIMMUN S-specific IgG and EUROIMMUN NCP-specific IgG kits.

## References

[1] Singh D and Yi SV. On the origin and evolution of SARS-CoV-2. Exp Mol Med, 2021. 53(4): p. 537–547.

[2] World Health Organization. COVID-19 Weekly Epidemiological Update. Edition 72. 28 December 2021. https://www.who.int/publications/m/item/weekly-epidemiological-update-on-covid-19---28-december-2021 [accessed 8 August 2022].

[3] WHO Director-General’s opening remarks at the media briefing on COVID-19 - 11 March 2020. https://www.who.int/director-general/speeches/detail/who-director-general-s-opening-remarks-at-the-media-briefing-on-covid-19---11-march-2020 [accessed 30th June 2021].

[4] Adepoju P. Nigeria responds to COVID-19; first case detected in sub-Saharan Africa. Nat Med, 2020. 26(4):444–448.

[5] Frost I, Osena G., Craig J, Hauck S, Kalanxhi E, Gatalo O, et al. COVID-19 in West Africa: National Projections of Total and Severe Infections Under Different Lockdown Scenarios. Center for Disease Dynamics, Economics & Policy. Department of Emergency Medicine, Johns Hopkins School of Medicine. 2020. https://cddep.org/wp-content/uploads/2020/07/West-Africa.pdf [accessed 6 July 2021].

[6] Villar J, Ariff S, Gunier RB, Thiruvengadam R, Rauch S, Kholin A, et al. Maternal and Neonatal Morbidity and Mortality Among Pregnant Women With and Without COVID-19 Infection: The INTERCOVID Multinational Cohort Study. JAMA Pediatr. 2021; 175(8):817–826.

[7] Median age of the population of Africa from 2000 to 2030. Statista. https://www.statista.com/statistics/1226158/median-age-of-the-population-of-africa/#:~:text=In%202022%2C%20the%20median%20age,it%20was%20around%2017%20years. [accessed 2 February 2023].

[8] Czeisler ME, Marynak K, Clarke KEN, Salah Z, Shakya I,Thierry JM, et al. Delay or Avoidance of Medical Care Because of COVID-19-Related Concerns - United States. June 2020. MMWR Morb Mortal Wkly Rep. 2020;69(36):1250–1257.

[9] Lewis HC, Ware H, Whelan M, Subissi L, Li Z, Ma X, Nardone A, et al. SARS-CoV-2 infection in Africa: a systematic review and meta-analysis of standardised seroprevalence studies, from January 2020 to December 2021. BMJ Glob Health. 2022;7(8).

[10] Kribi S, Toure F, Mendes A, Sanou S, Some A, Aminou AM, et al. Multicountry study of SARS-CoV-2 and associated risk factors among healthcare workers in Côte d’Ivoire, Burkina Faso and South Africa. Transactions of The Royal Society of Tropical Medicine and Hygiene. 2023;117(3):179–188.

[11] Tso FY, Lidenge SJ, Pena PB, Clegg AA, Ngowi JR, Mwaiselage J, et al. High prevalence of pre-existing serological cross-reactivity against severe acute respiratory syndrome coronavirus-2 (SARS-CoV-2) in sub-Saharan Africa. Int J Infect Dis. 2021;102:577–583.

[12] Lapidus S, Liu F, Casanovas-Massana A, Dai Y, Huck JD, Lucas, et al. Plasmodium infection is associated with cross-reactive antibodies to carbohydrate epitopes on the SARS-CoV-2 Spike protein. Sci Rep. 2022; 12(1):22175.

[13] World Health Organisation. 26 May 2020. Population-based age-stratified seroepidemiological investigation protocol for coronavirus 2019 (COVID-19) infection. Version 2.0. https://www.who.int/publications/i/item/WHO-2019-nCoV-Seroepidemiology-2020.2 [accessed 24 March 2023].

[14] Nkuba Ndaye A, Hoxha A, Madinga J, Marien J, Peeters M, Leendertz FH, et al. Challenges in interpreting SARS-CoV-2 serological results in African countries. Lancet Glob Health. 2021; 9(5):e588–e589.

[15] Our World in Data. Historical estimates of population, combined with the projected population to 2100 based on UN’s medium variant scenario. https://ourworldindata.org/grapher/population-past-future?tab=chart&country=GMB [accessed 26 July 2022].

[16] von Dadelszen, P, Flint-O’Kane M, Poston L, Craik R, Russell D, Tribe RM, et al. The PRECISE (PREgnancy Care Integrating translational Science, Everywhere) Network’s first protocol: deep phenotyping in three sub-Saharan African countries. Reprod Health. 2020;17 Suppl 1:51.

[17] Sanneh B, Sanneh S, Lareef-Jah S, Darboe B, Dibba LL, Manjang LF, et al. Practice of preventive measures and vaccine hesitance for COVID 19 among households in The Gambia, 2021: Study protocol. PLoS One. 2022; 17(8):e0270304.

[18] The Gambia COVID-19 Outbreak Situational Report No. 452. Epidemiology and Disease Control Unit, Ministry of Health, The Gambia. https://www.moh.gov.gm/wp-content/uploads/2022/09/GMB-COVID-19-Situational-Report-452_2022_10th_24th_September_2022.pdf [accessed 11 October 2022].

[19] Kanteh A, Jallow HS, Manneh J, Sanyang B, Kujabi MA, Ndure SL, et al. Genomic epidemiology of SARS-CoV-2 infections in The Gambia: an analysis of routinely collected surveillance data between March, 2020, and January, 2022. Lancet Glob Health. 2023;11(3):e414–e424.

[20] Abatan B, Agboghoroma O, Akemoke F, Antonio M, Awokola B, Bittaye M, et al. Intense and Mild First Epidemic Wave of Coronavirus Disease, The Gambia. Emerg Infect Dis. 2021;27(8):2064–2072.

[21] The Gambia COVID-19 Outbreak Situational Report No. 413. Epidemiology and Disease Control Unit, Ministry of Health, The Gambia. https://www.moh.gov.gm/wp-content/uploads/2021/12/GMB-COVID-19-Situational-Report_2021_25_26th_December_No-413.pdf [accessed 26 July 2022].

[22] The Gambia Bureau of Statistics. Distribution of Population by Gender and LGA. IHS 2015/2016. https://www.gbosdata.org/topics/population-and-demography/distribution-of-population-by-gender-and-lga [accessed 27 July 2022].

[23] WANTAI SARS-CoV-2 Ab ELISA. Ref WS-1096. Beijing Wantai Biological Pharmacy Enterprise Co., Ltd. No. 31 Kexueyuan Road, Changping District, Beijing, 102206 China. https://www.fda.gov/media/140929/download [accessed 27 July 2022].

[24] EUROIMMUN US. Anti-SARS-CoV-2 ELISA (IgG) Instruction for use. 1 Bloomfield Ave Mountain Lakes, New Jersey 07046. https://cdnmedia.eurofins.com/eurofinsus/media/1711222/ei_2606g_a_us_c01_igg_ce.pdf [accessed 27 July 2022].

[25] EUROIMMUN a PerkinElmer company. Anti-SARS-CoV-2 NCP ELISA (IgG) EI 2606-9601-2 G. Medizinische Labordiagnostika AG. Seekamp 31. 23560 Lubeck Germany. https://www.coronavirus-diagnostics.com/documents/Indications/Infections/Coronavirus/EI_2606_D_UK_C.pdf [accessed 12 August 2022].

[26] StataCorp. 2021. Stata Statistical Software: Release 17. College Station, TX: StataCorp LLC.

[27] Mohammed NI, Mackenzie G, Ezeani E, Sidibeh M, Jammeh L, Sarwar G, et al. Quantifying excess mortality during the COVID-19 pandemic in 2020 in The Gambia: a time-series analysis of three health and demographic surveillance systems. Int J Infect Dis. 2022; 128:61–68.

[28] Talla C, Loucoubar C, Roka JL, Barry A, Ndiaye S, Diarra M, et al. Seroprevalence of anti-SARS-CoV-2 antibodies in Senegal: a national population-based cross-sectional survey, between October and November 2020. IJID Reg. 2022;3:117–125.

[29] Konu YR, Conde S, Gbeasor-Komlanvi F, Sadio AJ, Tchankoni MK, Anani J, et al. SARS-CoV-2 antibody seroprevalence in Togo: a national cross-sectional household survey, May-June, 2021. BMC Public Health. 2022;22(1)2294.

[30] Emmerich P, Murawski C, Ehmen C, von Possel R, Pekarek N, Oestereich L, et al. Limited specificity of commercially available SARS-CoV-2 IgG ELISAs in serum samples of African origin. Trop Med Int Health. 2021;26(6)621–631.

[31] Navaratnam AMD, Shrotri M, Nguyen V, Braithwaite I, Beale S, Byrne TE, et al. Nucleocapsid and spike antibody responses following virologically confirmed SARS-CoV-2 infection: an observational analysis in the Virus Watch community cohort. Int J Infect Dis. 2022;123:104–111.

[32] Alfego D, Sullivan A, Poirier B, Williams J, Adcock D, Letovsky S. A population-based analysis of the longevity of SARS-CoV-2 antibody seropositivity in the United States. EClinicalMedicine. 2021;36100902.

[33] Downe S, Finlayson K, Tuncalp LJ, Metin Gulmezoglu A. What matters to women: a systematic scoping review to identify the processes and outcomes of antenatal care provision that are important to healthy pregnant women. BJOG. 2016;123(4):529–39.

