## Supplementary Table 1 for "SARS-CoV-2 seroprevalence in pregnant women during the first three COVID-19 waves in The Gambia"

**Supplementary Table 1. Characteristics of study women at each period**

| Characteristic | Pre-pandemic1  N=112 | Pre-pandemic2  N=73 | Post-wave1  N=209 | Post-wave2  N=208 | Post-wave3  N=201 |
| --- | --- | --- | --- | --- | --- |
| N:  Urban  Rural | 87  25 | 68  5 | 77  132 | 97  111 | 75  126 |
| Age, years  median (IQR) | 26 (21.5, 32) | 28 (24, 32) | 26 (21, 32) | 26 (21, 31) | 27 (22, 34) |
| Ethnicity % (n)  Mandinka  Wolof  Fula  Others | 44.6 (50)  46.4 (52)  5.4 (6)  3.6 (4) | 35.6 (26)  45.2 (33)  16.4 (12)  2.7 (2) | 41.1 (86)  26.8 (56)  29.2 (61)  2.9 (6) | 41.8 (87)  34.1 (71)  23.6 (49)  0.5 (1) | 39.8 (80)  21.9 (44)  33.8 (68)  4.5 (9) |
| Household size  median (IQR) | 9 (6, 9) | 8 (5, 9) | 9 (8, 9) | 9 (6, 9) | 9 (8, 9) |
| Parity  median (IQR) | 3 (2, 5) | 4 (2, 5) | 2 (1, 5) | 3 (1, 5) | 3 (1, 5) |
