## Supplementary Table 2 for "SARS-CoV-2 seroprevalence in pregnant women during the first three COVID-19 waves in The Gambia"

**Supplementary Table 2. Prevalence of SARS-CoV-2 S-protein IgG and N-protein IgG at each post-wave study period and comparison of prevalence for S-protein and N-protein IgGs within each post-wave period.**

| Period | S-protein IgG | | NCP-protein IgG | | S-protein IgG vs NCP-protein IgG | | | |
| --- | --- | --- | --- | --- | --- | --- | --- | --- |
|  | Positive | Prevalence (95% CI) | Positive | Prevalence (95% CI) | PR  (95% CI) | p-value | PD  (95% CI) | p-value |
| Post-Wave1  N=209 | 34 | 16.3  (11.9, 21.9) | 27 | 12.9  (9.0, 18.1) | 1.3  (0.6,2.5) | 1 | 3.3  (-6.8, 13.5) | 1 |
| Post-Wave2  N=208 | 82 | 39.4  (33, 46.2) | 54 | 26  (20.5, 32.3) | 1.5  (1.0,2.3) | 0.061 | 13.5  (0.1, 26.8) | **0.047** |
| Post-Wave3  N=211 | 150 | 74.6  (68.2, 80.1) | 104 | 51.7  (44.9, 58.6) | 1.4  (1.1, 1.8) | **<0.001** | 22.9  (9.2, 36.6) | **<0.001** |
