## Supplementary figures and images for "SARS-CoV-2 seroprevalence in pregnant women during the first three COVID-19 waves in The Gambia"

### Supplementary Figure 1

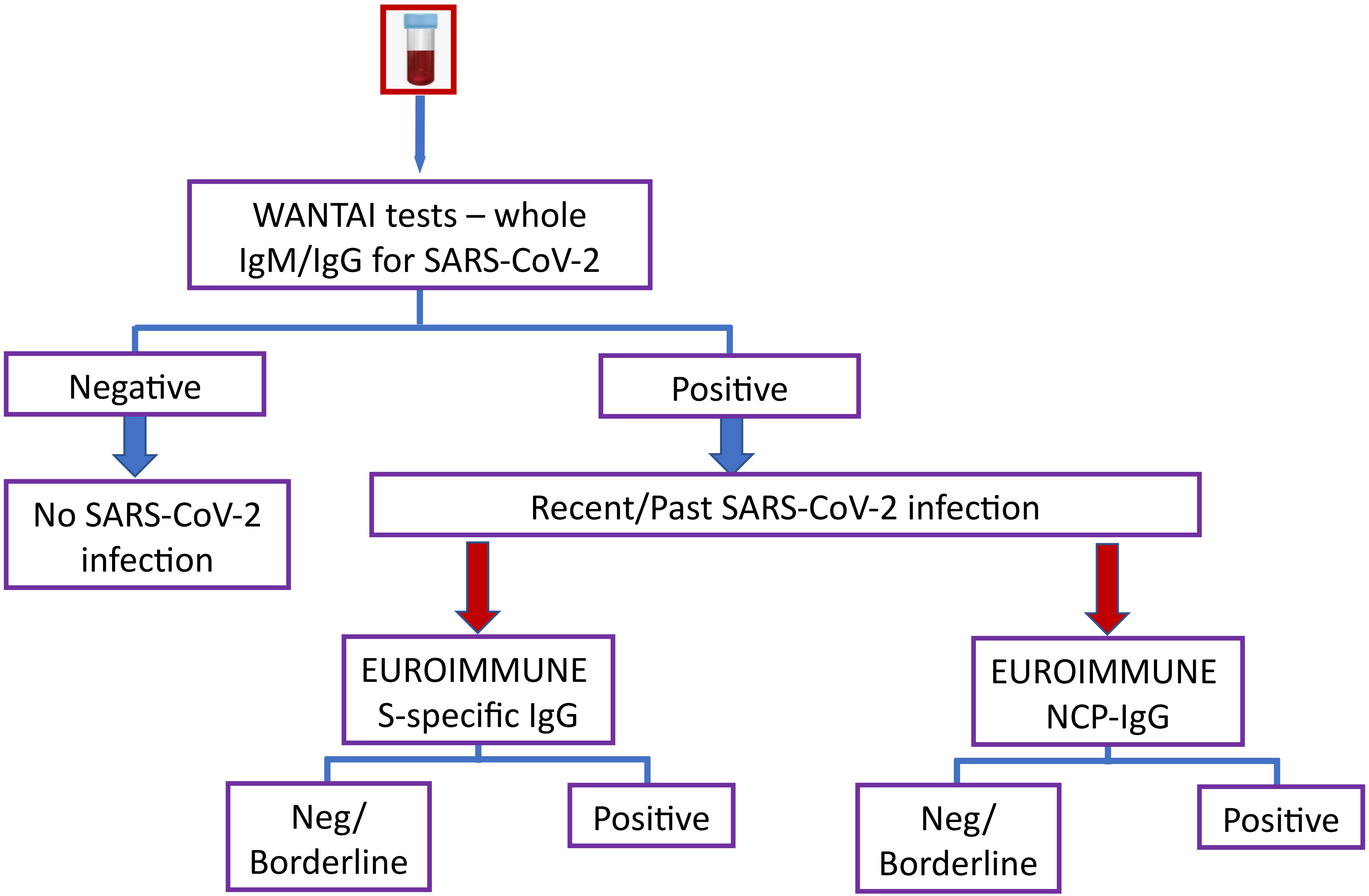
